# Prediction of Homologous Recombination Deficiency from Oncomine Comprehensive Assay Plus Correlating with SOPHiA DDM HRD Solution

**DOI:** 10.1101/2023.08.09.23293743

**Authors:** Jun Kang, Kiyong Na, Haeyoun Kang, Uiju Cho, Sun Young Kwon, Sohyun Hwang, Ahwon Lee, The Molecular Pathology Study Group of Korean Society of Pathologists

**Affiliations:** Department of Hospital Pathology, Seoul St. Mary’s Hospital, College of Medicine, The Catholic University of Korea, Seoul, Korea; Department of Pathology, Kyung Hee University College of Medicine, Kyung Hee University Hospital, Seoul, Republic of Korea; Department of Pathology, CHA Bundang Medical Center, CHA University, Seongnam, Korea; Department of Pathology, St. Vincent’s Hospital, College of Medicine, The Catholic University of Korea, Seoul, Korea; Department of pathology, Dongsan Hospital, School of Medicine, Keimyung University; CHA Future Medicine Research Institute, CHA Bundang Medical Center, Seongnam, Korea; Cancer Research Institute, The Catholic University of Korea, Seoul, Korea

## Abstract

**Objective:** Poly(ADP-ribose) polymerase (PARP) inhibitors are used for targeted therapy for ovarian cancer with homologous recombination deficiency (HRD). In this study, we aimed to develop a homologous recombination deficiency prediction model to predict the genomic integrity (GI) index of the SOPHiA DDM HRD Solution from the Oncomine Comprehensive Assay (OCA) Plus. We also tried to a find cut-off value of the genomic instability metric (GIM) of the OCA Plus that correlates with the GI index of the SOPHiA DDM HRD Solution.

**Methods:** We included 87 cases with high-grade ovarian serous carcinoma from five tertiary referral hospitals in Republic of Korea. We developed an HRD prediction model to predict the GI index of the SOPHiA DDM HRD Solution. As predictor variables in the model, we used the HRD score, which included percent loss of heterozygosity (%LOH), percent telomeric allelic imbalance (%TAI), percent large-scale state transitions (%LST), and the genomic instability metric (GIM), provided by the OCA Plus. To build the model, we employed a penalized logistic regression technique.

**Results:** The final model equation is −21.77 + 0.200 × GIM + 0.102 × %LOH + 0.037 × %TAI + 0.261 × %LST. To improve the performance of the prediction model, we added a borderline result category to the GI results. Cases with predicted values between −3 and 3 were classified as borderline. The accuracy of our HRD status prediction model was 0.947 for the training set and 0.958 for the test set. The accuracy of HRD status using GIM with a cut-off value of 16 was 0.911.

**Conclusions:** The Oncomine Comprehensive Assay Plus provides a reliable biomarker for homologous recombination deficiency.

**What is already known on this topic**

The Oncomine Comprehensive Assay Plus is a targeted next-generation sequencing assay designed to detect genetic alterations in solid tumors. It has not been validated as a biomarker for PARP inhibitor response through clinical trials or a concordance test with a Food and Drug Administration–approved homologous recombination deficiency test.

**What this study adds**

This study introduces a predictive model for homologous recombination deficiency using data from the Oncomine Comprehensive Assay Plus, which correlates with the SOPHiA DDM HRD Solution. The study provides evidence that the Oncomine Comprehensive Assay Plus is a reliable biomarker for homologous recombination deficiency.

**How this study might affect research, practice or policy**

The strong agreement between the Oncomine Comprehensive Assay Plus and the SOPHiA DDM HRD Solution suppports its potential as a biomarker for predicting PARP inhibitor response. This finding could have implications for PARP inhibitor-related research, clinical practice, and regarding their use in clinical trials.

## INTRODUCTION

Homologous recombination repair (HRR) is a DNA repair mechanism that restores DNA double-strand breaks (DSBs) in cells. This mechanism is essential for maintaining genomic stability and preventing the accumulation of DNA damage that can lead to mutations and other genetic alterations. When HRR is impaired, such as through mutations in genes involved in this repair pathway, it can lead to a condition known as homologous recombination deficiency (HRD). HRD has been found to be associated with an increased risk of developing certain types of cancer, including ovarian, breast, and prostate cancer^1 2^. The genes most commonly associated with HRD are *BRCA1* and *BRCA2*, which are tumor suppressor genes that play a critical role in HRR^3^. Mutations in these genes can impair the HRR pathway, leading to an increased risk of developing cancer. Other genes involved in HRR, such as *PALB2* and *RAD51*, have also been linked to HRD and an increased cancer risk^4 5^.

Poly(ADP-ribose) polymerase (PARP) inhibitors such as olaparib and talazoparib are a type of targeted therapy that work by inhibiting the function of poly(ADP-ribose) polymerase 1 (PARP1), an enzyme that is involved in the repair of single-strand DNA breaks (SSBs). PARP inhibitors have shown clinical efficacy in BRCA1/2 mutant ovarian cancer, breast cancer, and prostate cancer^6–9^. These drugs have demonstrated promising results in clinical trials and have been approved by regulatory agencies for the treatment of certain types of cancer.

HRD can lead to abnormal DSB repair and result in genomic scars, which are large-scale genomic alterations that can be quantified by counting the number of occurrences. There are several types of genomic scars associated with HRD, including large-scale loss of heterozygosity (LOH)^10^, telomere allelic imbalance (TAI)^11^, and large-scale state transitions (LST)^12^. The HRD score is a quantification of these genomic scars and is used to identify patients who may benefit from treatment with PARP inhibitors^13 14^. The HRD score is calculated based on the occurrence of these genomic scars. These tests, such as the Myriad myChoice CDx and FoundationOne CDx tests, have been approved by regulatory agencies as companion diagnostics for PARP inhibitor treatment in patients with ovarian and prostate cancer^6 8 15 16^.

The SOPHiA DDM HRD Solution is a HRD test that identifies HRR mutations through targeted sequencing and measures genomic instability (GI) through a combination of low-pass whole-genome sequencing and a deep-learning algorithm^17^. The GI index is a measure of genomic stability of the SOPHiA DDM HRD Solution. This index is based on the analysis of the genome-wide patterns of copy number variations (CNVs) and is used to determine the level of GI in a tumor sample. A high GI index indicates a high level of GI and is associated with HRD tumors^17^.

The Oncomine Comprehensive Assay (OCA) Plus is a targeted next-generation sequencing (NGS) assay designed to detect genetic alterations in solid tumors. The HRD score provided by the OCA includes (1) percent LOH (%LOH), which estimates the fraction of the genome with LOH identified using genomic segmentation; (2) percent TAI (%TAI), which estimates the fraction of the genome with allelic imbalance or unequal contribution from the two alleles in the telomeres identified using genomic segmentation; and (3) percent LST (%LST), which estimates the fraction of the genome with unequal copy numbers in adjacent segments identified using genomic segmentation. These values range from 0 to 100. The genomic instability metric (GIM) is a proprietary measurement that quantifies genomic scarring associated with HRD.

In this study, we aimed to develop an HRD prediction model to predict the GI index of the SOPHiA DDM HRD Solution from the OCA Plus. We also tried to find a cut-off value of the GIM of the OCA Plus that correlates with the GI index of the SOPHiA DDM HRD Solution.

## METHODS

### Sample Collection

We included 87 cases of high-grade ovarian serous carcinoma from five tertiary referral hospitals in Republic of Korea. All cases had been tested with OCA Plus NGS panel for clinical purpose at the hospitals where patients were treated. We excluded the cases that failed to analyze HRD scores provided by the OCA Plus. In all cases, we confirmed the clinical information and tissue diagnosis and we selected paraffin blocks for the SOPHiA DDM HRD Solution. We cut all formalin-fixed paraffin-embedded (FFPE) tissue samples to a thickness of 5 μm. We sent 10 sections to the institution in the Republic of Korea that performs the SOPHiA DDM HRD Solution.

### Oncomine Comprehensive Assay Plus

We extracted genomic DNA by using the Maxwell RSC DNA FFPE Kit (Promega, Madison, WI, USA) in accordance with the instructions provided by the manufacturer. We determined the DNA concentration by using the Qubit ds DNA High-Sensitive Assay kit (Thermo Fisher Scientific, Waltham, MA, USA) on the Qubit fluorometer (Thermo Fisher Scientific).

We performed all manual library preparation by using the OCA Plus system (Thermo Fisher Scientific), following the manufacturer’s instructions. We conducted the multiplex polymerase chain reaction (PCR) amplification with an approximate DNA concentration of 20 ng. Prior to PCR amplification, we carried out the deamination reaction in the OCA Plus by using Uracil-DNA Glycosylase, heat labile (Thermo Fisher Scientific).

For sequencing, we loaded the prepared libraries onto Ion 550 Chips (Thermo Fisher Scientific) according to the manufacturer’s instructions and processed them using the Ion Chef System. We used the Ion S5 XL Sequencer (Thermo Fisher Scientific) for sequencing. We aligned the data to the human genome assembly 19, which served as the standard reference genome in the Ion Reporter Software (v. 5.18) (Thermo Fisher Scientific). Hospital B utilized the customized variability control informatics baseline (VCIB) for copy number analysis. The GIM was obtained from the Ion Reporter Software (v. 5.20).

### Genomic Instability Score Prediction Modeling

We developed an HRD prediction model that aimed to predict the GI index of the SOPHiA DDM HRD Solution. The training set consisted of cases from hospital A, while the test set comprised cases from the other hospitals. The predictor variables used in the model were the HRD score, which included %LOH, %TAI, %LST, and the GIM, provided by the OCA Plus. To build the model, we employed a penalized logistic regression technique. We selected the model through repeated fivefold cross-validation on a grid of hyperparameters: λ (10^−5^, 10^−4^, 10^−3^, 10^−2^, and 10^−1^) and α (0.0, 0.25, 0.5, 0.75, and 1.0).

### Assessing Model Performance

We estimated the performance of the prediction based on the area under the curve of the receiver operating characteristic curve (AUROC) for the GI status and the HRD status. We considered the GI status to be positive when the GI index exceeded 0. On the other hand, we considered the HRD status to be positive if there was a BRCA1/2 pathogenic variant or if the GI status was positive. We conducted the modeling and assessment of model performance using the tidymodels and glmnet R packages. A flowchart of the study is presented in Supplemental Figure 1.

In accordance with the journal’s guidelines, we will provide our data for independent analysis by a selected team by the Editorial Team for the purposes of additional data analysis or for the reproducibility of this study in other centers if such is requested.

### Research Ethics and Patient Consent

The study was approved by the Institutional Review Board of the CHA Bundang Medical Center, CHA University (2023-01-010-001) and the Catholic University of Seoul Saint Mary’s Hospital (KC18TNSI0361), where this study was organized.

## RESULTS

### Patients

The average age of the patients was 59.3 years. The majority of patients had advanced disease based on the The International Federation of Gynecology and Obstetrics (FIGO) stage. Hospital A contributed the most cases (55, accounting for 63.2% of the total). There were no significant statistical differences in patient age and the FIGO stage between the train set and the test set, as shown in Table 1.

**Table 1.** Cases summary.

|  | Overall | Training set | Test set | p value |
| --- | --- | --- | --- | --- |
| Number of patients | 87 | 55 | 32 |  |
| Age (mean (SD)) | 59.3 (10.3) | 59.2 (10.5) | 59.4 (10.2) | 0.946 |
| FIGO stage (%) |  |  |  | 0.875 |
| 1 | 7 (8.1) | 5 (9.3) | 2 (6.2) |  |
| 2 | 6 (7.0) | 3 (5.6) | 3 (9.4) |  |
| 3 | 63 (73.3) | 40 (74.1) | 23 (71.9) |  |
| 4 | 10 (11.6) | 6 (11.1) | 4 (12.5) |  |
| Institution (%) |  |  |  |  |
| Hospital A | 55 (63.2) | 55 (100.0) | 0 (0.0) |  |
| Hospital B | 15 (17.2) | 0 (0.0) | 15 (46.9) |  |
| Hospital C | 10 (11.5) | 0 (0.0) | 10 (31.2) |  |
| Hospital D | 4 (4.6) | 0 (0.0) | 4 (12.5) |  |
| Hospital E | 3 (3.4) | 0 (0.0) | 3 (9.4) |  |

SD: standard deviation, FIGO: The International Federation of Gynecology and Obstetrics

### SOPHiA DDM HRD Solution

The HRD status was positive in 56 cases (64.4%), negative in 23 cases (26.4%), and undetermined in 8 cases (9.2%). The GI status was positive in 50 cases (57.5%), negative in 27 cases (31.0%), and undetermined in 10 cases (11.5%). The BRCA status was positive in 28 cases (32.2%), negative in 45 cases (51.7%), and undetermined in 14 cases (16.1%).

### Oncomine Comprehensive Assay Plus Sequencing

The mean of the average base coverage was 2469.64. The mean of the median absolute pairwise difference (MAPD) was 0.24. The MAPD is a metric that measures read coverage noise detected across all amplicons in a panel. A higher MAPD typically indicates lower coverage uniformity, which can result in missed or erroneous CNV calls. The quality control parameter metrics are summarized in Online Supplemental Table 1.

### BRCA1/2 Pathogenic Variants

The concordance rate for BRCA1/2 pathogenic variants between the SOPHiA DDM HRD Solution and the OCA Plus was 95.9%. The discordant cases included two frameshift variants at homopolymer sequences. The OCA Plus pipeline filtered out these pathogenic variants due to an unusual prediction filter that measured the amount of strand bias according to the manufacturer’s specifications. These two frameshift variants were restored by modifying the parameter of the unusual prediction filter. The other discordant variant was a long deletion. The long deletion could not be detected because it spanned across the ends of amplicons. The pathogenic variants found in BRCA1/2 are listed in Online Supplemental Table 2.

### Selecting Model and Performance Estimation

After excluding cases without a GI index from the SOPHiA DDM HRD Solution, the training set and test set consisted of 51 and 26 cases, respectively. The model with a penalty of 0.1 and a mixture of 1 (Lasso regression) demonstrated the best performance in terms of the AUROC in a repeated fivefold cross-validation (Supplemental Figure 2). We fit the final model with the selected hyperparameters using the entire training set. The final model equation is −21.77 + 0.200 × GIM + 0.102 × LOH(%) + 0.037 × TAI(%) + 0.261 × LST (%). To improve the performance of the prediction model, we added a borderline result category to the GI results. We classified cases with predicted values between −3 and 3 as borderline (Figure 1). The accuracy of our HRD status prediction model was 0.947 for the training set and 0.958 for the test set. Detailed performance metrics are summarized in Table 2.

**Figure 1.**
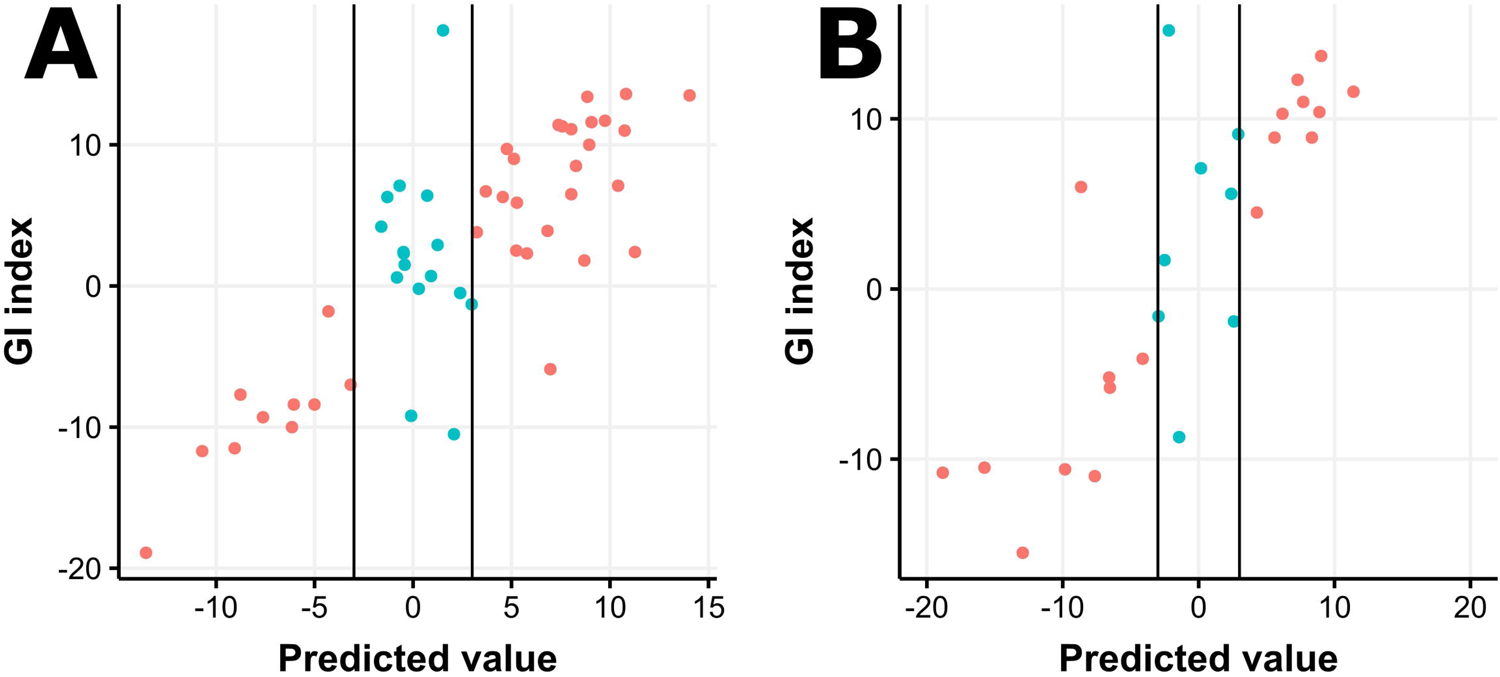
Performance of the homologous recombination deficiency (HRD) prediction model. The training set (A) and the test set (B) are shown. The black vertical lines represent borderline cut-off values (−3 and 3).

**Table 2.** Performance metrics.

| HRD test | N | Accuracy | Sensitivity | Specificity | Positive<br>Predictive<br>Value | Negative<br>Predictive<br>Value | F1 score |
| --- | --- | --- | --- | --- | --- | --- | --- |
| HRD result (Train) | 38 | 0.947 95% CI ( 0.823 - 0.994 ) | 0.964 | 0.900 | 0.964 | 0.900 | 0.964 |
| HRD result (Test) | 24 | 0.958 95% CI ( 0.789 - 0.999 ) | 0.938 | 1.000 | 1.000 | 0.889 | 0.968 |
| HRD result (GIM) | 79 | 0.911 95% CI ( 0.826 - 0.964 ) | 0.964 | 0.783 | 0.915 | 0.900 | 0.939 |
| GI result (Train) | 33 | 0.97 95% CI ( 0.842 - 0.999 ) | 1.000 | 0.909 | 0.957 | 1.000 | 0.978 |
| GI result (Test) | 23 | 0.87 95% CI ( 0.664 - 0.972 ) | 0.786 | 1.000 | 1.000 | 0.750 | 0.880 |
| GI result (GIM) | 77 | 0.883 95% CI ( 0.79 - 0.945 ) | 0.960 | 0.741 | 0.873 | 0.909 | 0.914 |

HRD: homologous recombination deficiency, HRD result (Train/Test): prediction model performance for HRD result of SOPHiA DDM
HRD Solution, GI: Genomic Integrity, GI Result (Train/Test): prediction model performance for GI result of SOPHiA DDM HRD Solution
F1 score: harmonic mean of the precision and recall, GIM: Genomic Instability Metric, HRD result/GI result (GIM): prediction performance
of GIM for HRD result/GI result of SOPHiA DDM HRD Solution

### Genomic Instability Metric

The AUROC for the GI status of SOPHiA DDM HRD Solution was 0.887 (Figure 2A). We set the positive cut-off value at 16 (Figure 2B). The accuracy of the HRD status using the GIM with a cut-off value of 16 was 0.911. Detailed performance metrics are summarized in Table 4.

**Figure 2.**
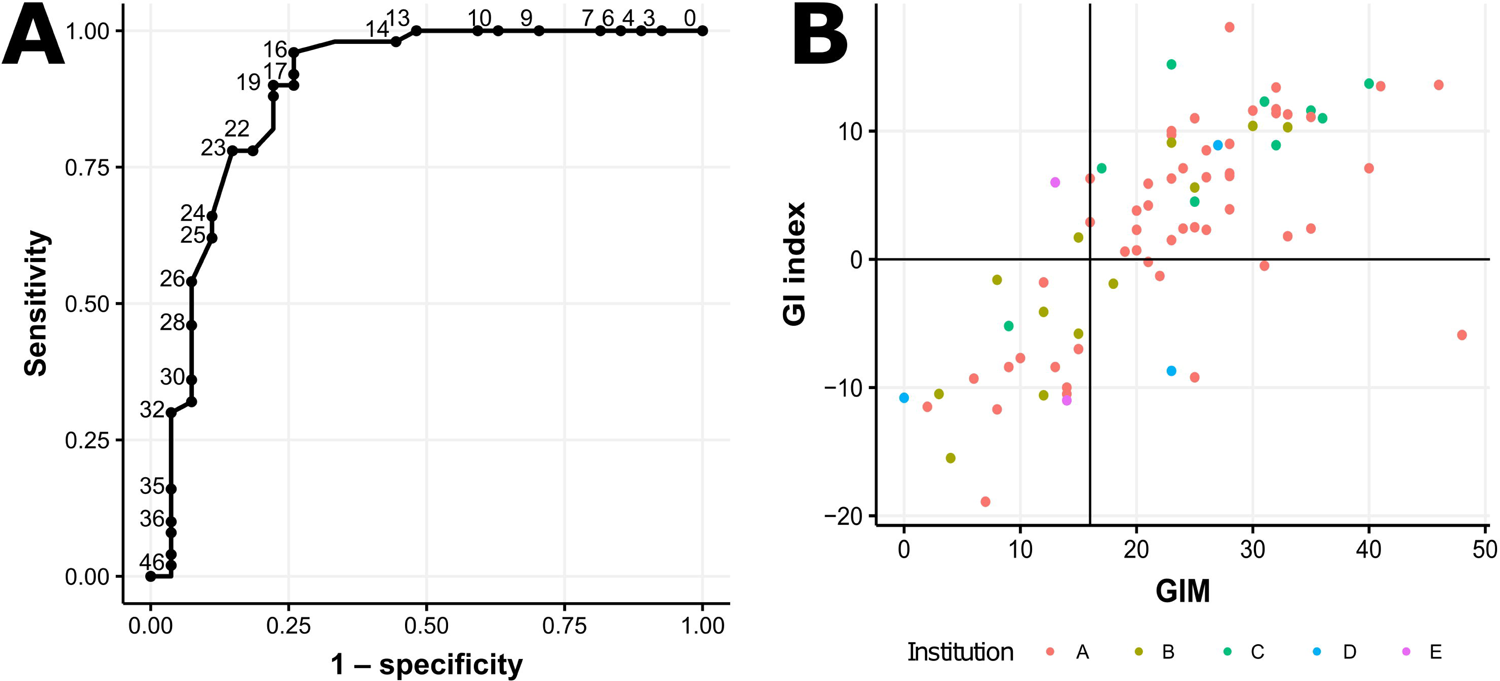
Receiver operating characteristic curve (ROC) for the genomic instability (GI) status of the SOPHiA DDM HRD Solution (A). The threshold (cut-off value) was set at 16. The black vertical line indicates a genomic instability metric (GIM) of 16. The black horizontal line indicates a GI index of 0.

## DISCUSSION

### Summary of Main Results

In this study, we developed a penalized linear regression model using the OCA Plus, which showed a high concordance rate with the SOPHiA DDM HRD Solution. We also observed that the GIM of the OCA Plus demonstrated high accuracy compared with the SOPHiA DDM HRD Solution. Despite being independently developed by different manufacturers, the GIM of the OCA Plus and the GI index of the SOPHiA DDM HRD Solution exhibited a high concordance rate. These findings suggest that both tests capture the same tumor characteristic, namely genomic alteration associated with HRD. When two different tests yield the same results, it reinforces the certainty of the results. It also indicates that both tests are reliable and reproducible.

### Results in the Context of Published Literature

It is important to note that neither the OCA Plus nor the SOPHiA DDM HRD Solution has been validated as a biomarker for PARP inhibitor response through clinical trials. Both the OCA Plus and the SOPHiA DDM HRD Solution require clinical validation through a clinical trial or a concordance test with a Food and Drug Administration–approved HRD test.

The SOPHiA DDM HRD Solution exhibited a considerable rate of failure, resulting in an undetermined result. This failure rate is similar to that of the myChoice HRD Plus assay^15^. Additionally, the OCA Plus fails to analyze HRD scores, and our prediction model relies on the OCA Plus HRD scores. Our prediction model includes a borderline category, which does not definitively determine the GI status. However, the GIM also demonstrated high accuracy without the need for a borderline category.

### Strengths and Weaknesses

In three cases (4%), the OCA Plus failed to detect BRCA1/2 pathogenic variants. Two of these were c.2175del [chr13:32910667del] and c.3503dup [chr17:41244048dup] and were filtered out due to the application of a filter related to strand bias originating from homopolymer sequences. These false negatives were restored by modifying the filter parameter. The remaining one is the c.2593_2621del [chr17:41244928_41244956del] mutation, a 26 base pair deletion located within the overlapping regions of the OCA Plus amplicons. This type of long deletion seems to interfere with the generation of libraries containing both amplicons carrying this mutation, resulting in the absence of sequencing reads. It has been observed that the coverage depth of these two amplicons is relatively low compared with the adjacent amplicon positions. It is anticipated that detecting this mutation with the OCA Plus would be challenging. Therefore, interpretation of BRCA1/2 status results should consider the limitations of the test. These two cases had high GI and were classified as HRD positive.

Because the NGS study was not conducted on all patients with high-grade ovarian serous carcinoma, the patients included in this study may exhibit bias. However, the rates of BRCA1/2 pathogenic variant presence and positive HRD status are similar to those reported in a clinical trial of ovarian high-grade serous carcinoma using the myChoice HRD Plus assay (Myriad Genetic Laboratories).

We developed the penalized linear regression model by using a small training set and validated it with a small test set. This approach may lead to a model that is either too simplistic and underfits the data or too complex and overfits the data.

### Implications for Practice and Future Research

The OCA Plus offers several advantages compared with HRD-specific tests. It enables comprehensive analysis of genetic alterations, including single nucleotide variants (SNVs), insertions and deletions (indels), CNVs, structural variations, the tumor mutation burden, mismatch repair deficiency, and microsatellite instability. This broad coverage enhances the ability to identify potential targeted treatments. The high accuracy between the OCA Plus and the SOPHiA DDM HRD Solution supports its potential as a biomarker for predicting the PARP inhibitor response and its application in clinical trials for PARP inhibitors. Additionally, our study provides a cut-off value for the GIM of the OCA Plus that correlates with the SOPHiA DDM HRD Solution, with a high accuracy of 0.911.

## CONCLUSIONS

This study presents a homologous recombination deficiency prediction model from the Oncomine Comprehensive Assay Plus that correlates with the SOPHiA DDM HRD Solution. this study provides evidences that The Oncomine Comprehensive Assay Plus provides reliable biomarkers for homologous recombination.

## Data Availability

All data produced in the present study are available upon reasonable request to the authors

## Acknowledgement

The authors thank Taeeun Kim for providing data acquisition.

