## Supplemental Material for "Prediction of Homologous Recombination Deficiency from Oncomine Comprehensive Assay Plus Correlating with SOPHiA DDM HRD Solution"

Supplemental Table 1. The quality control parameter metrics of Oncomine Comprehensive Assay Plus

|  | Overall | training | test | p value |
| --- | --- | --- | --- | --- |
| Number of cases | 87 | 55 | 32 |  |
| Tumor cellularity (%) (mean (SD)) | 67.52 (17.71) | 69.51 (15.16) | 64.34 (21.02) | 0.198 |
| Average base coverage depth (mean (SD)) | 2469.64 (462.08) | 2348.31 (481.15) | 2678.19 (343.58) | 0.001 |
| MAPD (mean (SD)) | 0.24 (0.06) | 0.24 (0.06) | 0.25 (0.06) | 0.359 |

SD: standard deviation, MAPD: median absolute pairwise difference

Supplemental Table 2. List of BRCA1/2 pathogenic variants

| Gene | Nucleotide change (SOPHiA) | Nucleotide change (OCA Plus) | Amino acid change (SOPHiA) | Amino acid change (OCA Plus) |
| --- | --- | --- | --- | --- |
| BRCA1 | c.4186C>T | c.4186C>T | p.Gln1396* | p.Gln1396Ter |
| BRCA2 | c.7322del | c.7322del | p.Gly244Alafs*26 | p.Gly244AlafsTer26 |
| BRCA1 | c.3627dup | c.3627dup | p.Glu1210Argfs*9 | p.Glu1210ArgfsTer9 |
| BRCA1 | c.922_924delinsT | c.922_924delAGCinsT | p.Ser308* | p.S308X |
| BRCA2 | c.2175del | Not detected | p.Val726Phefs*4 | Not detected |
| BRCA1 | c.4318G>T | c.4318G>T | p.Glu1440* | p.Glu1440Ter |
| BRCA1 | c.2593_2621del | Not detected | p.Lys865Serfs*28 | Not detected |
| BRCA2 | c.5576_5579del | c.5576_5579delTTAA | p.Ile1859Lysfs*3 | p.Ile1859LysfsTer3 |
| BRCA2 | c.6239T>G | c.6239T>G | p.Leu2080* | p.Leu2080Ter |
| BRCA1 | c.3593T>A | c.3593T>A | p.Leu1198* | p.Leu1198Ter |
| BRCA1 | c.922_924delinsT | c.922_924delAGCinsT | p.Ser308* | p.Ser308Ter |
| BRCA2 | c.5576_5579del | c.5576_5579delTTAA | p.Ile1859Lysfs*3 | p.Ile1859LysfsTer3 |
| BRCA1 | c.3503dup | Not detected | p.Asn1168Lysfs*2 | Not detected |
| BRCA1 | c.4484+1G>T | c.4484+1G>T | NA | NA |
| BRCA1 | c.3442del | c.3442delG | p.Glu1148Argfs*7 | p.Glu1148ArgfsTer7 |
| BRCA2 | c.8143A>T | c.8143A>T | p.Lys2715* | p.Lys2715Ter |
| BRCA1 | c.4485-1G>T | c.4485-1G>T | NA | NA |
| BRCA1 | c.3296del | c.3296delC | p.Pro1099Leufs*10 | p.Pro1099LeufsTer10 |
| BRCA1 | c.922_924delinsT | c.922_924delAGCinsT | p.Ser308* | p.Ser308Ter |
| BRCA1 | c.3875dup | c.3875_3876insC | p.Ala1293Cysfs*2 | p.Ala1293CysfsTer2 |
| BRCA2 | c.6449_6450del | c.6449_6450delAA | p.Lys2150Serfs*25 | p.Lys2150SerfsTer25 |
| BRCA1 | c.2062A>T | c.390C>A | p.Thr688Ser | p.Tyr130Ter |
| BRCA1 | c.4629del | c.4629delG | p.Pro1544Hisfs*4 | p.Pro1544HisfsTer4 |
| BRCA1 | c.4986+5G>A | c.4986+5G>A | NA | NA |
| BRCA2 | c.7008-1G>T | c.7008-1G>T | NA | NA |
| BRCA2 | c.5576_5579del | c.5576_5579delTTAA | p.Ile1859Lysfs*3 | p.Ile1859LysfsTer3 |
| BRCA2 | c.7516C>T | c.7516C>T | p.Gln2506* | p.Gln2506Ter |
| BRCA2 | c.3599_3600del | c.3599_3600delGT | p.Cys1200* | p.Cys1200Ter |

OCA: Oncomine Comprehensive Assay
