## Supplementary figures and images for "Prediction of Homologous Recombination Deficiency from Oncomine Comprehensive Assay Plus Correlating with SOPHiA DDM HRD Solution"

### Supplemental Figure 1

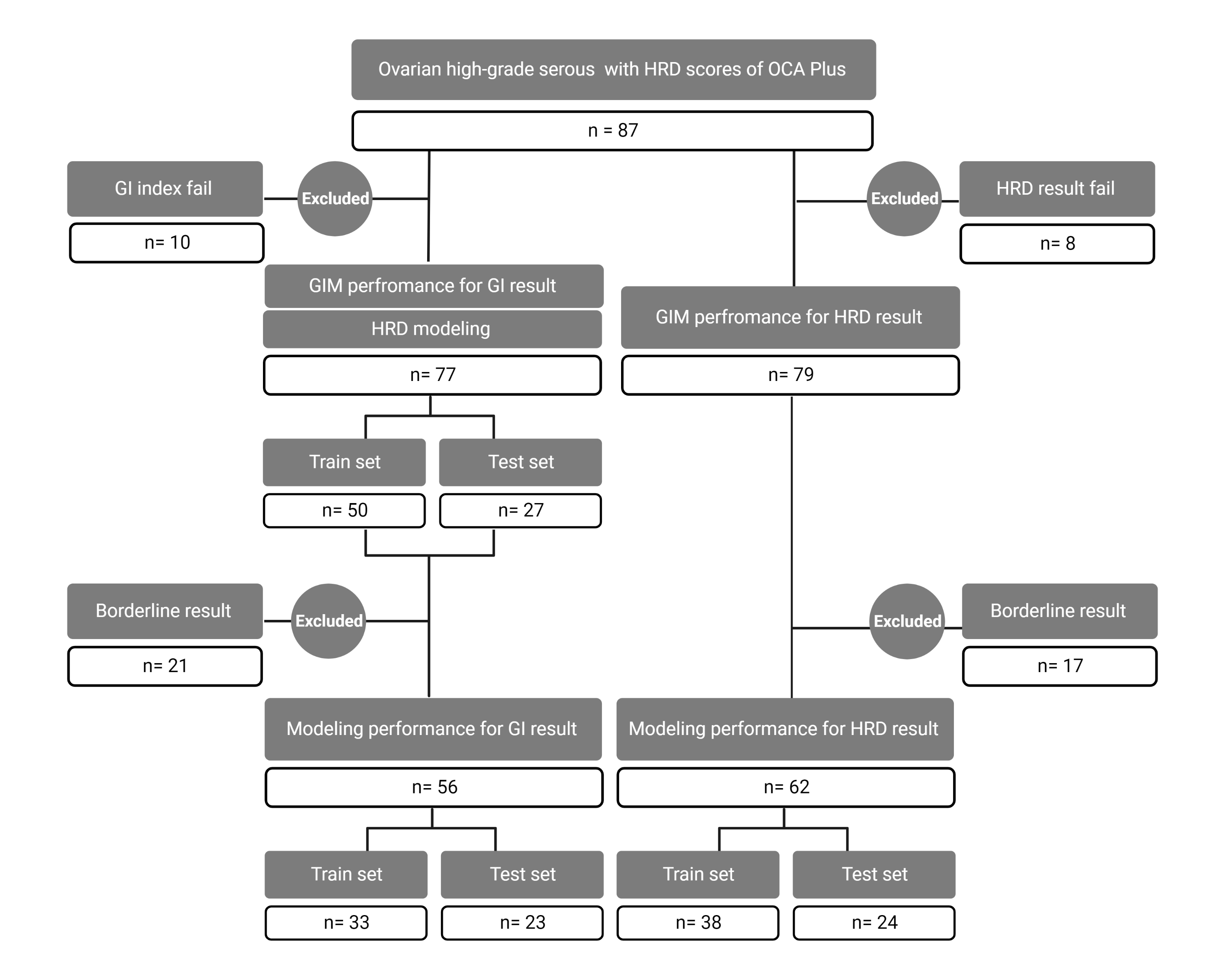

### Supplemental Figure 2

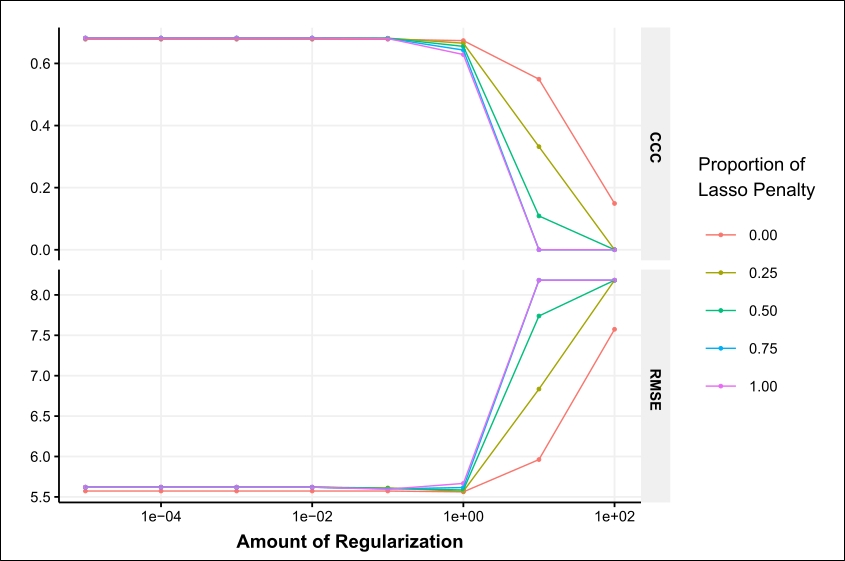
