## Supplemental Figure Legends for "Prediction of Homologous Recombination Deficiency from Oncomine Comprehensive Assay Plus Correlating with SOPHiA DDM HRD Solution"

Supplemental Figure 1. Flowchart of the study.

Supplemental Figure 2. Hyperparameter tuning and performance assessment in repeated 5-fold cross-validation on a grid of hyper-parameters. The x-axis is a penalty scaling parameter: $\lambda$(${10}^{-5}$, ${10}^{-4}$, ${10}^{-3}$, ${10}^{-2}$, ${10}^{-1}$, ${10}^{0}$), color is mixture hyperparameter of penalty function: $\alpha$ (0.0, 0.25, 0.5, 0.75, 1.0). CCC: concordance correlation coefficient, RMSE: root mean squared error
